# Assessment of the humoral response to the homologous Gam-COVID-Vac (Sputnik V) or heterologous Sputnik V/mRNA-1273 (Moderna) vaccination against SARS-CoV-2 in dialysis patients

**DOI:** 10.1101/2022.05.13.22275049

**Authors:** Gustavo Laham, Alfredo P. Martínez, Wanda Rojas Gimenez, Lucas Amaya, Anabel Abib, Natalia Echegoyen, Carlos Díaz, Alicia Lucero, Antonella Martelli, Cristina Videla, Karin Neukam, Federico A. Di Lello

## Abstract

**Introduction:** The humoral response to vaccines is the most used tool to evaluate the protection against SARS-CoV-2 infection. Dialysis patients are a high-risk population and have a reduced immune response to vaccination.

**Objective:** To assess the humoral response to homologous Gam-COVID-Vac (Sputnik V) and heterologous Sputnik V/mRNA-1273 (Moderna) vaccination in dialysis patients.

**Methods:** SARS-CoV-2 anti-spike IgG (RBD) concentration was estimated 3-16 weeks after complete vaccination. Reactogenicity was evaluated until day 7 by patients’
s self-reported side events.

**Results:** 107 participants were enrolled [n=84 homologous (SpV/SpV), n=23 heterologous (SpV/Mod)]. Median (IQR) age was 64 (50-75) years old and 79 (73.8%) were male. Additionally, 19 (22.6%) of the SpV/SpV and 4 (17.4%) of the SpV/Mod group had a prior confirmed SARS-CoV-2 infection (p=0.589). In the overall population, 103 patients reached seroconversion (96.3%). Anti-S-RBD IgG median titers (IQR) were higher in the heterologous [1222 (288-5680) BAU/mL] than in the homologous scheme [447 (100-1551) BAU/mL], p=0.022. In a linear model adjusted for age and gender, previous SARS-COV-2 infection (B: 1944.3; CI95: 1136.2-2753.4; p<0.001), and SpV/Mod vaccination scheme (B: 1241.5; CI95: 420.39-2062.6; p=0.003) were independently associated with anti-S-RBD levels. Finally, a higher frequency of adverse effects was associated with the heterologous scheme, although they were well tolerated by all individuals.

**Conclusion:** The present study provides evidence that the homologous SpV/SpV and heterologous SpV/Mod schemes showed good efficacy and safety under dialysis conditions. These results could be useful for future vaccination strategies, especially aimed at this risk group.

## INTRODUCTION

Patients under dialysis are at increased risk for severe coronavirus infectious disease 2019 (COVID-19) caused by the severe acute respiratory syndrome coronavirus 2 (SARS-CoV-2) than the general population, with reported mortality rates of up to 28.3% [1-3-]. This situation mainly obeys to a considerably higher mean patients’ age (approximately 65 years old) and the frequent presence of comorbid conditions such as obesity, diabetes, and high blood pressure that are linked with a more severe COVID-19 course. Fortunately, effective vaccination schemes against SARS-CoV-2 inducing reduction of both infection and the risk of severe COVID-19 have been rapidly developed [4-7-]. Moreover, when facing first dose serious adverse effects or dose supply shortcomings, introduced heterologous vaccination schemes have shown promising results [8-12-]. Nevertheless, clinical trials do not provide information about vaccine efficacy in dialysis populations and possible differences in immunogenicity among SARS-CoV-2 vaccination schemes are poorly understood due to their novelty. Particularly, the homologous Gam-COVID-VAC (Sputnik V) and the heterologous vaccination scheme including Sputnik V as a prime dose and mRNA-1273 (Moderna) as a booster dose have been barely studied since Sputnik V is not approved in all countries and its implementation suffers geographic limitations. In Argentina, the National Administration of Medicines, Food, and Medical Technology (ANMAT) has approved Sputnik V use; however, due to the second dose shortage, mRNA-1273 (Moderna) has been frequently administered as the booster dose. In this scenario, the present study aimed to assess the humoral response to homologous Sputnik V and heterologous Sputnik V/Moderna vaccination in dialysis patients.

## MATERIAL AND METHODS

### Study design and population

From March to October 2021, subjects who attended for hemodialysis at the Centro de Educación Médica e Investigaciones Clínicas “Norberto Quirno” (CEMIC), Buenos Aires, Argentina, were included in this prospective cohort study. Inclusion criteria were i) patients who had received Sputnik V prime immunization, ii) they had received a boost dose of either Sputnik V (SpV/SpV) or Moderna (SpV/Mod) vaccines within 18 weeks post-prime dose, and iii) they presented to monitor their humoral immune response three weeks after the boost dose. The vaccination scheme depended on dose availability and the prioritization of risk populations as established by the Argentine Ministry of Health.

### Immunogenicity

Binding IgG antibodies against the spike (S) receptor-binding domain (RBD) of SARS-CoV-2 (anti-S-RBD) concentration was assessed at 3-16 weeks after boost. Anti-S-RBD antibodies were quantified using the Abbott Diagnostics SARS-CoV-2 IgG II Quant chemiluminescent microparticle immunoassay (CMIA) on an Architect i2000 SR and an Alinity I analyzer (Abbott Diagnostics, Abbott Park, Illinois, USA). To standardize the results to WHO binding antibody units (BAU), a correction factor for Abbott arbitrary units (AU) was applied where 1 BAU/mL equals 0.142 AU, as previously established by Abbott with the WHO international standard NIBSC 20–136 [13-. Following the manufacturer’s recommendations, samples were considered reactive for anti-S-RBD when titers were above 50 AU/mL (7.2 BAU/mL). An 80% protective effect (PROT-80) against symptomatic SARS-CoV-2 infection was assumed when anti-S-RBD titers were 506 BAU/ml or higher [14-.

### Reactogenicity

All patients were invited to complete an online questionnaire to report all possible post-boost vaccination adverse events and required medical assistance. The intensity of adverse effects was graded as mild, moderate, and severe depending on the interference with daily activities.

### Statistical analysis

Descriptive statistics and univariate analyses were performed. The outcome variable was the anti-S-RBD titer at least three weeks after the boost dose. Differences in anti-S-RBD levels and PROT-80 according to demographic and clinical parameters were evaluated. Categorical variables were expressed as numbers (percentage) and analyzed using the Chi-square test or the Fisher’s test. The student’s t-test and the Mann-Whitney *U* test were used to compare independent continuous variables, expressed as median (interquartile range, IQR). For related continuous variables, the Wilcoxon signed-rank test was applied. Those factors associated with anti-S-RBD levels with a p<0.2 in the univariate analysis, were evaluated in a generalized linear model adjusted for age and sex. Likewise, multivariate logistic regression models were developed to identify factors associated with PROT-80. Adjusted odds ratios (AOR) with their corresponding 95% confidence intervals (CI95) were calculated. Statistical analyses were carried out using the SPSS statistical software package release 23.0 (IBM SPSS Inc., Chicago, IL, USA).

### Ethical Approval and Informed Consent

The study was designed and performed according to the Helsinki declaration and all participants gave their written informed consent (Study protocol EX-2021-06438339-UBA DME#SSA_FFYB, Ethics committee of the Facultad de Farmacia y Bioquímica, Universidad de Buenos Aires).

## RESULTS

### Study population

A total of 107 subjects were included in the study, 84 (78.5%) received SpV/SpV, and the remaining 23 (21.5%) the SpV/Mod scheme. Seventy-nine (73.8%) participants were male, and the median (IQR) age was 64 (50-75) years old. Overall, median time intervals were 91 (77-116) days from prime to boost dose (ΔP-B) and 32 (24-47) days from the boost dose to the anti-S-RBD IgG serological determination (ΔB-antiSRBD). Eighty-four (78.5%) individuals were *naïve* to SARS-CoV-2 infection at the time of prime vaccination. Then, 19 (22.6%) of those who received the SpV/SpV scheme and 4 (17.4%) who received the SpV/Mod one had a prior confirmed SARS-CoV-2 infection (p=0.589). Table 1 shows detailed characteristics of the study population.

**Table 1.** Population epidemiological characteristics (n=107).

| Characteristic | Total, n=107 |
| --- | --- |
| Age* (years) | 64 (50-75) |
| Male gender, n (%) | 79 (73.8) |
| Primary kidney diseases |  |
| Diabetes, n (%) | 15 (14) |
| High blood pressure, n (%) | 13 (12.1) |
| Polycystic kidney disease, n (%) | 8 (7.5) |
| Glomerular diseases, n (%) | 10 (9.3) |
| Other, n (%) | 23 (21.5) |
| Unknown, n (%) | 38 (35.6) |
| Time on dialysis* (years) | 4 (2-7) |
| Weekly total Kt/V median* | 1.5 (1.3-1.9) |
| Laboratory |  |
| Hemoglobin* (g/dL) | 10.8 (9.7-11.6) |
| Albumin* (g/L) | 4.1 (3.7-4.3) |
| C-reactive protein* (mg/dL) | 4.6 (1.4-10.0) |
| Transferrin saturation* | 28 (23-36) |
| Body mass index (BMI)* (kg/m<sup>2</sup>) | 25.3 (22.4-27.9) |
| Obesity (BMI >30 kg/m<sup>2</sup>) | 13 (12.1) |

\*Median (interquartile range).

### Immunogenicity

In the overall population, anti-S-RBD IgG was reactive in 103 (96.3%) persons, 80 (95.2%) immunized with the SpV/SpV vaccine, and 23 (100%) with the SpV/Mod one (p=0.286). Median (IQR) anti-S-RBD titers were 42.5 (4-1297) BAU/mL after the first dose and 502 (110-1993) BAU/mL after the boost dose (p<0.001). In participants with a confirmed SARS-CoV-2 infection before vaccination receiving the SpV/SpV scheme, the humoral response as measured by anti-S-RBD levels was 6.5-fold higher than that observed in *naïve* ones. Similarly, people with a confirmed SARS-CoV-2 infection before the SpV/Mod scheme administration presented 11-fold higher anti-S-RBD levels when compared to participants without prior infection (Figure 1). Anti-S-RBD levels according to epidemiological and clinical parameters are shown in Table 2. Previous COVID-19 (B: 1944.3; CI95: 1136.2-2753.4; p<0.001) and SpV/Mod vaccination scheme (B:1241.5; CI95: 420.39-2062.6; p=0.003) were independently associated with anti-S-RBD levels in a linear model adjusted for age (B:−1.297; CI05: −23.20-20.61; p=0.907), sex (B:−371.92; 95CI −1131.5-387.66; p=0.334), ΔP-B (B:54.911; 95CI: −36.641-146.46; p=0.237), and ΔB-antiSRB (B:-7.828; CI95: −107.04-91.383; p=0.876).

**Figure 1.**
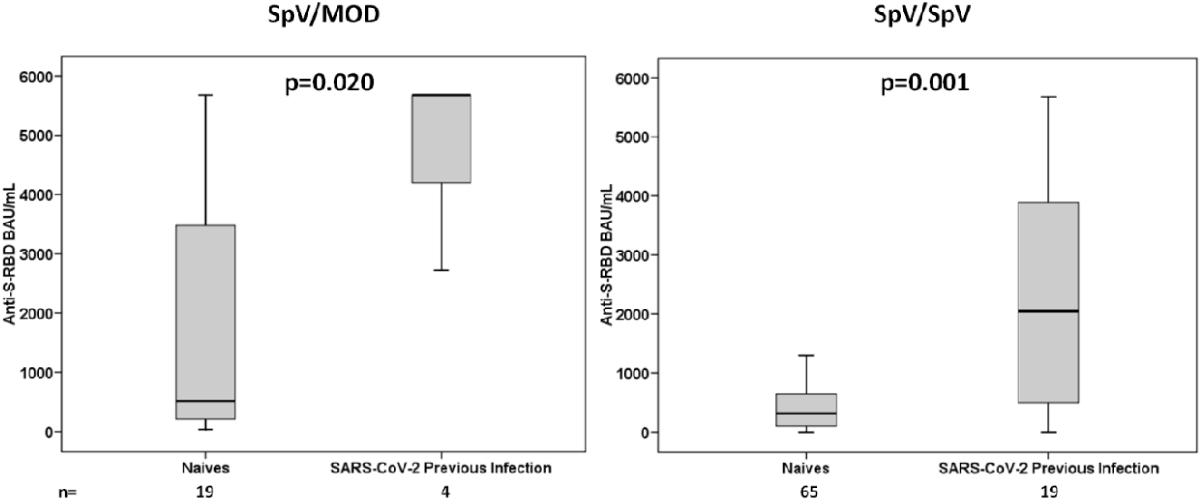
Anti-S-RBD IgG levels for homologous (SpV/SpV) and heterologous (SpV/Mod) vaccination schemes in subjects who had (A) or had not (B) a confirmed SARS-CoV-2 infection prior to immunization.

**Table 2.** Anti-S-RBD IgG and PROT-80 according to epidemiological and vaccination-specific parameters (N=107)

| Parameter | Anti-S-RBD |  |  | PROT-80 |  |
| --- | --- | --- | --- | --- | --- |
|  | N | (BAU/mL)* | *p* | N (%) | *p* |
| Age |  |  |  |  |  |
| 30-49 years | 25 | 605 (285-1396) |  | 17 (68.0%) |  |
| 50-60 years | 19 | 501 (88-914) |  | 7 (36.8%) |  |
| >60 years | 63 | 342 (100-2873) | 0.770 | 29 (46%) | 0.084 |
| Gender |  |  |  |  |  |
| Female | 28 | 431 (105-2419) |  | 14 (50.0%) |  |
| Male | 79 | 502 (110-1740) | 0.859 | 39 (49.4%) | 0.954 |
| Time on dialysis |  |  |  |  |  |
| ≤3 years | 42 | 345 (100-1551) |  | 18 (42.8) |  |
| ≥4 years | 65 | 555 (112-2634) | 0.346 | 35 (53.8) | 0.267 |
| Vaccination scheme |  |  |  |  |  |
| SpV/SpV | 84 | 447 (100-1442) |  | 38 (45.2%) |  |
| SpV/Mod | 23 | 1222 (288-5680) | 0.022 | 15 (65.2%) | 0.090 |
| Prior SARS-CoV-2 infection |  |  |  |  |  |
| No | 84 | 339 (99-911) |  | 35 (41.7%) |  |
| Yes | 23 | 2872 (508-5680) | <0.001 | 18 (78.3%) | 0.002 |
| ΔP-B |  |  |  |  |  |
| 3-10 weeks | 19 | 288 (94-612) |  | 7 (36.8%) |  |
| 11-18 weeks | 88 | 518 (117-2761) | 0.141 | 46 (52.3%) | 0.312 |
| ΔB-antiSRBD |  |  |  |  |  |
| 3-4 weeks | 51 | 443 (114-1303) |  | 23 (45.1%) |  |
| 5-8 weeks | 41 | 584 (105-3852) |  | 24 (58.5%) |  |
| 9-12 weeks | 9 | 502 (126-2436) |  | 4 (44.4%) |  |
| 13-16 weeks | 6 | 120 (53-608) | 0.187 | 2 (33.3%) | 0.484 |

SpV = Sputnik V, Mod = Moderna
\*Median (interquartile range); time intervals from prime to boost ( ΔP-B) and from boost to anti-
S-RBD IgG determination ( ΔB-antiSRBD).
Weekly total Kt/V median, Hemoglobin, Albumin, C-reactive protein, Transferrin saturation, and
Body mass index showed no association with Anti-S-RBD or PROT-80.

A total of 53 (49.5%) individuals achieved PROT-80. Among participants without prior COVID-19 who received SpV/SpV or SpV/Mod and those with confirmed COVID-19 who received the homologous and the heterologous schemes, PROT-80 rates were 36.9%, 57.9%, 73.7%, and 100% (p_linear association_<0.001), respectively. Corresponding values according to epidemiological and clinical parameters are displayed in Table 2. In the multivariate analysis, an independent association with PROT-80 was observed for prior COVID-19 (AOR: 8.663; CI95: 2.541-29.540; p=0.001), a heterologous vaccination scheme (AOR: 3.767; CI95: 1.229-10.921; p=0.015) and age (AOR: 0.962; CI95: 0.933-0.992; p=0.012), in a model adjusted for these parameters, as well as sex (AOR: 0.883; CI95: 0.331-2.361; p=0.805).

### Reactogenicity

The homologous and heterologous immunization schemes were well tolerated, and no medical assistance or potentially fatal events were reported. Adverse events, including local and systemic symptoms, were higher for SpV/Mod (47.6%) than for SpV/SpV (23.7%) schemes, p=0.031. In general, the most frequent systemic adverse events were fatigue (9.9%), myalgia (5.9%), and fever (2.0%). No patients reported headaches, chills, nausea/vomiting, arthralgia, or diarrhea. The heterologous vaccine scheme tends to induce more systemic adverse effects than the homologous one (28.6% vs 15.0%, p=0.148). Regarding local adverse events, pain at the injection site was reported by 11 patients (10.9%) and tended to be more frequent for the heterologous scheme than for the homologous one (19.0% vs 8.8%, p=0.178). Figure 2 shows the reactogenicity by adverse effects (local and systemic) according to the vaccination scheme.

**Figure 2.**
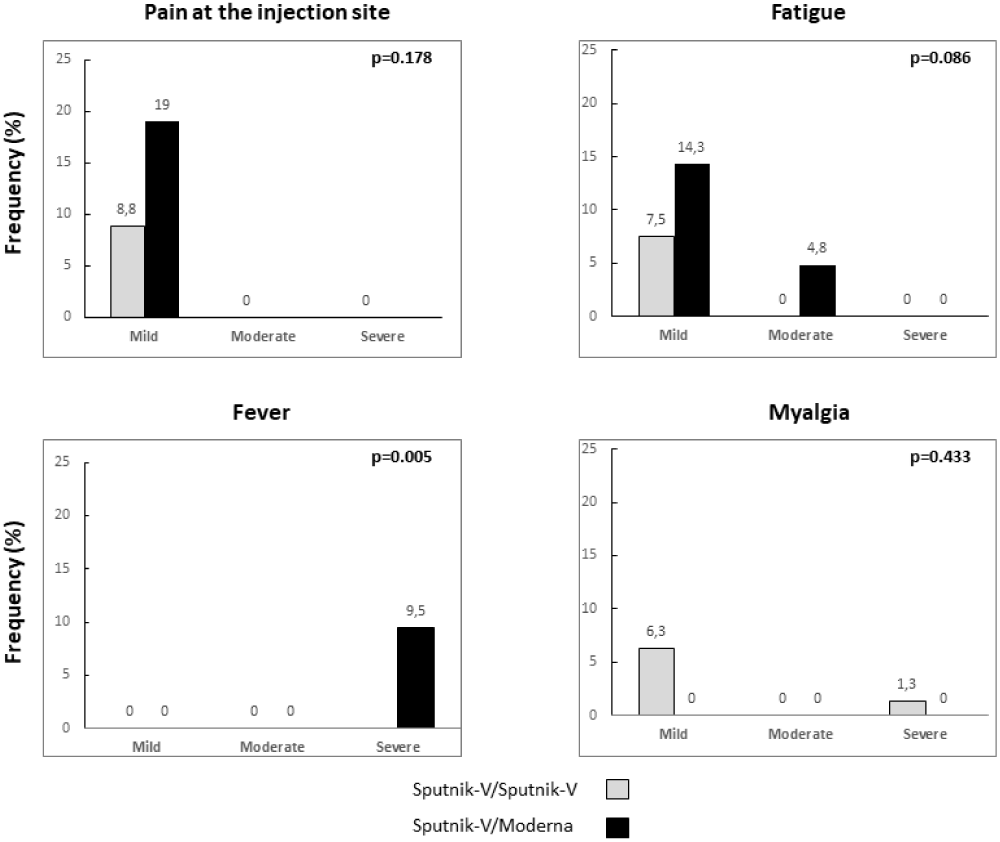
Reactogenicity: frequency of local and systemic adverse effects as reported by the participants for the homologous (SpV/SpV) and heterologous (SpV/Mod) schemes, classified by severity.

## DISCUSSION

The present work shows that the implementation of SpV/SpV or SpV/Mod vaccination schemes against SARS-CoV-2 in patients under dialysis was effective leading to a seroconversion rate of 96.3%. Overall, the heterologous scheme showed an anti-S-RBD level almost 3-fold higher than the homologous one. Moreover, both vaccination schemes were well tolerated, and no medical assistance was required.

To our knowledge, this is the first study on the heterologous scheme including Sputnik V and Moderna vaccines in *naïve* and previously SARS-CoV-2 infected patients under dialysis conditions. Additionally, the study allowed results comparison with the homologous SpV/SpV scheme. The seroconversion rate of our population under dialysis was similar to that reported by previous studies showing values between 80-98% [15-20-]. Rosa-Diez (2021) detected a 98% seroconversion rate in a study including 102 dialysis patients vaccinated with the Sputnik V scheme [16-]. On the other hand, studies performed on patients under dialysis conditions immunized with Moderna vaccines have shown seroconversion rates ranging between 95% and 97.9% [15-,18-,21-. Recent evidence suggested that the mRNA vaccine has a better humoral response when compared to adenovirus-based vaccines [8-,9-,22-24-. In fact, the higher anti-S-RBD titer achieved in this study with the SpV/Mod group agrees with previous works performed on the general population [12-26-]. In this context, our results support the use of Sputnik V or its combination with the Moderna vaccine as an alternative for dialysis patients. However, additional studies are necessary to assess the cellular response since the combination of vaccines appears to enhance the characteristic immune response generated by each vaccination scheme [22-,27-].

Results from our study showed the development of a strong humoral response in the dialysis setting, even when some works have shown that patients under dialysis present a lower seroconversion rate and anti-S-RBD titers than healthy controls [28-].

Regarding PROT-80, almost 50% of the studied population achieved this threshold while heterologous scheme and prior SARS-CoV-2 infection were associated with higher PROT-80 proportions. Furthermore, the multivariate analysis showed that the 80% vaccine efficacy was not associated with age, gender, ΔP-B, and ΔB-antiSRBD. Obtained results were in accordance with our previous work performed on general population immunized with the SpV/SpV and SpV/MOD schemes [12-].

In the present study, there were no anti-S-RBD IgG significant differences between genders. These findings are consistent with several studies showing that gender seems not to influence anti-S-RBD IgG titers achieved with both analyzed vaccine schemes [28-32-].

No anti-S-RBD IgG difference according to the age of dialyzed patients was observed in our sample. Available data on the relationship between age and response to vaccines are scarce and controversial [30-,32-34-]; importantly, older ages were associated with lower rates of PROT-80. More studies on this issue are warranted as most reports analyzing the age-associated responses are conducted in healthy populations [30-,32-34-].

Regarding previous infection with SARS-CoV-2, a significant correlation was found between higher anti-S-RBD IgG titers and a confirmed past infection for both the SpV/SpV and SpV/MOD schemes, with a notable difference (11 times) for the heterologous scheme. This finding is in accordance with our previously reported values for the same vaccination schemes [12-], for the sputnik V homologous scheme [29-30-] and for RNAm vaccines [35-].

Additionally, the current study found that the body mass index, time on dialysis, weekly total Kt/V, as well as other evaluated laboratory parameters showed no association with Anti-S-RBD or PROT-80 levels. There are previous contradictory results regarding this issue. While some authors, as we did, could not find an association between these variables and the anti-S-RBD IgG titers [36-, other studies suggest that less comorbidity presence leads to a higher anti-S-RBD IgG titer [37-38-].

There is little data on SpV/SpV or SpV/Mod reactogenicity under dialysis conditions and this work offers a contribution to the understanding of SARS-CoV-2 vaccination in this setting. In our sample, adverse events including local and systemic symptoms were higher in dialyzed patients who received the SpV/Mod scheme as we have previously described for the general population [12-]. In agreement with other studies, fatigue and myalgia were the most frequent systemic reactions [12-,39-]. The higher trend in the rate of adverse events in the SpV/Mod group was expected since it is well proven that stronger side effects were associated with mRNA vaccines in the general population [11-,12-27-40-,41-. It is important to note that the frequency of adverse events was lower than that observed in the general population and no patients required medical support [12-]. However, the median age of the sample analyzed in the present work was higher and the effect of age on reactogenicity has already been described in previous studies [4-,5-,42-]. Moreover, in a Polewska’s (2021) study, adverse events were less frequently observed in dialyzed patients than in the age and sex-matched control group [39-].

This study has a limitation. It should be considered that serious adverse effects have been reported in a very low frequency and the small size of the analyzed sample might have influenced this aspect.

In conclusion, both analyzed vaccine schemes were immunogenic and showed a high seroconversion rate. In addition, a significant correlation was found between higher anti-S-RBD IgG titers and a confirmed prior infection with SARS-CoV-2 for both schemes. Moreover, the heterologous scheme was also associated with a better humoral response. Finally, local and systemic adverse effects were scarce and mostly mild, demonstrating that both schemes are safe and well-tolerated. These findings should promote patients on dialysis to receive these immunization schemes.

## Data Availability

All data produced in the present work are contained in the manuscript

## Contributors and authorship

**FAD, GL and KN:** Study concept and design, analysis, and interpretation of data, drafting of the manuscript and statistical analysis and study supervision.

**WRG, AA, NE, CD, AL, AM** and **CV:** Acquisition of data, critical revision of the manuscript for important intellectual content and final approval of the version to be submitted. **APM:** Acquisition of data, analysis and interpretation of data, critical revision of the manuscript for important intellectual content and final approval of the version to be submitted.

## ACKNOWLEDGMENT

FAD is a member of the National Research Council (CONICET) Research Career Program. K.N. is the recipient of a Miguel Servet contract by the Instituto de Salud Carlos III (grant number CPII18/00033). We would like to thank Mrs. Silvina Heisecke, from CEMIC-CONICET, for the copyediting of the manuscript

